# Wearable-Grade Lead Reduction Disproportionately Degrades ECG AI Performance in Elderly Patients: Evidence from PTB-XL and MIT-BIH

**DOI:** 10.64898/2026.06.12.26355537

**Authors:** Kabi Raj Tiruwa, Abhisan Ghimire

## Abstract

Consumer wearable devices increasingly use single-lead electrocardiograms (ECGs) for cardiac monitoring, but these signals contain substantially less spatial information than the clinical 12-lead standard. Whether this reduction disproportionately affects older adults, who often present with more complex cardiac conditions, remains poorly understood. In this study, we evaluated the impact of lead reduction on AI-ECG diagnostic performance across age groups. A 1D residual neural network was trained on 21,091 PTB-XL ECG recordings spanning five diagnostic superclasses and assessed using 12-, 6-, 2-, and 1-lead configurations. Under the full 12-lead setting, model accuracy declined from 84.5% in patients younger than 40 years to 66.2% in patients aged 75 years or older. Progressive lead reduction further widened this gap. Under the 1-lead configuration, accuracy decreased by 14.1 percentage points in the 75+ group but by only 0.4 percentage points in the **<40** group, representing an approximately 40-fold differential degradation confirmed by three independent statistical tests (all ***p* < 0.0001**). Older adults also exhibited greater multi-condition diagnostic complexity, providing a plausible explanation for their increased vulnerability to information loss. External validation on the MIT-BIH Arrhythmia Database confirmed cross-dataset model stability. These findings suggest that age-stratified performance reporting should be a minimum standard in wearable AI-ECG validation and regulatory assessment.

## 1 Introduction

Cardiovascular disease causes approximately 17.9 million deaths each year and accounts for 32% of global mortality [1, 2]. The standard 12-lead electrocardiogram (ECG) remains the primary non-invasive tool for cardiac screening [3]. Heart failure alone affects more than 64 million people worldwide, with incidence increasing as populations age, particularly among individuals older than 75 years [4]. However, conventional 12-lead ECG acquisition requires trained healthcare professionals and specialized clinical equipment, which limits its scalability for large community-based and remote screening programs.

Deep learning models applied to 12-lead ECGs can classify arrhythmias with performance comparable to trained cardiologists [5], diagnose multiple cardiac conditions automatically [6], detect hidden signs of atrial fibrillation even when the ECG shows normal sinus rhythm [7], and predict future onset of atrial fibrillation across large populations with strong performance (AUROC 0.83) [8]. Multi-label classification has also shown that multiple co-existing cardiac conditions can be identified simultaneously from a single ECG [9]. These advances have generated broad interest in AI-enabled ECG screening; however, all of these models were developed and validated using complete 12-lead ECG input.

Consumer wearable devices such as Apple Watch and Fitbit are now widely used and have regulatory approval for ECG recording [10, 11]. Large-scale studies, including the Apple Heart Study (n=419,297) [10] and Fitbit Heart Study (n=455,699) [12], showed that wearable devices can detect atrial fibrillation (AF) at population scale. Although AI models using single-lead wearable ECGs have shown promising results [13], these devices record far less information than clinical 12-lead ECG systems [14]. This raises an important question of whether AI models trained on full 12-lead ECGs can maintain performance with limited wearable data. While AI-ECG has been clinically validated in a prospective non-randomized interventional trial and shown to improve atrial fibrillation (AF) detection [15], age-specific effects of lead reduction remain unclear.

Despite growing evidence that demographic factors influence AI-ECG performance, studies have shown that prediction accuracy can vary by race, sex, and age [16]. AI models can also detect racial patterns from ECG signals, even though race itself is not a direct biological electrical marker [17], suggesting that social and clinical inequalities may shape these predictions [18]. Because many AI systems are trained on historical clinical data, they may also inherit existing healthcare disparities and demographic bias [19]. Prior work has further demonstrated demographic bias in ECG AI models, where performance may differ systematically across patient groups [20]. However, it remains unclear whether reducing ECG leads in wearable settings disproportionately affects diagnostic accuracy across age groups. As equitable AI deployment becomes a broader priority in medicine, this question requires systematic investigation.

External validation, where a model trained on one patient cohort is tested on a completely independent dataset with different hardware, sampling rates, and patient populations, is a critical but often overlooked step in ECG AI research [5, 6, 8]. It remains unclear whether models trained on PTB-XL using reduced leads (12, 6, 2, or 1) can generalize reliably to independent arrhythmia datasets. Additionally, whether older patients experience greater accuracy loss under lead reduction consistently across different datasets has not yet been systematically investigated.

We trained a 1D residual neural network on the PTB-XL dataset (n=21,091; 5 superclasses: Normal, MI, ST/T change, conduction disturbance, and hypertrophy) [21, 22]. The model was evaluated under four ECG lead configurations (12, 6, 2, and 1-lead) and across four age groups to measure performance differences. We studied the interaction between lead reduction and age, showing an approximately 40-fold differential degradation in accuracy. We further examined whether multi-condition diagnostic complexity explains this effect. For external validation, the 2-lead model was tested without retraining on the MIT-BIH Arrhythmia Database (n=46) [22, 23] to assess cross-dataset generalization.

## 2 Methods

### 2.1 Datasets

Two publicly available open-access datasets were used in this study: PTB-XL and MIT-BIH [21, 22]. PTB-XL is a large 12-lead ECG database (n=21,799; 100 Hz; 10-second recordings) collected at Leipzig University Hospital between 1989 and 1996 and distributed through PhysioNet [23, 24]. Before model training, 284 records with invalid age values (*>*100 years) were excluded, resulting in 21,091 ECGs with valid superclass labels. ECGs were grouped into five broad diagnostic superclasses derived from SCP annotations: NORM (normal ECG), MI (myocardial infarction), STTC (ST/T change), CD (conduction disturbance), and HYP (hypertrophy), providing standardized and clinically interpretable classification. Stratified 10-fold splitting was used to preserve class balance, with folds 1–8 for training (n=16,862), fold 9 for validation (n=2,104), and fold 10 as the held-out test set (n=2,125). The MIT-BIH Arrhythmia Database (n=48; 2-lead MLII and V5; 360 Hz) was used exclusively for external validation [22, 23]. Two records with invalid age data were excluded (final n=46). No MIT-BIH data were used for training. Both datasets are publicly available, deidentified, and open-licensed; therefore, no institutional ethics review was required [22].

### 2.2 Lead Configurations

Four lead configurations were evaluated in this study (Table 1). The 12-lead configuration represents the full hospital-grade ECG and serves as the clinical gold standard. The 6-lead configuration (limb leads I, II, III, aVR, aVL, and aVF) reflects standard clinical monitoring systems. The 2-lead configuration (Lead II + V5) matches the MIT-BIH acquisition format and common ambulatory monitoring devices. The 1-lead configuration (Lead II only) replicates the single-lead ECG recorded by consumer wearable devices such as the Apple Watch [15]. A separate model was trained independently for each lead configuration using identical training data and hyperparameters, ensuring that any observed performance differences were attributable only to lead count rather than differences in training methods or model optimization.

**Table 1.**
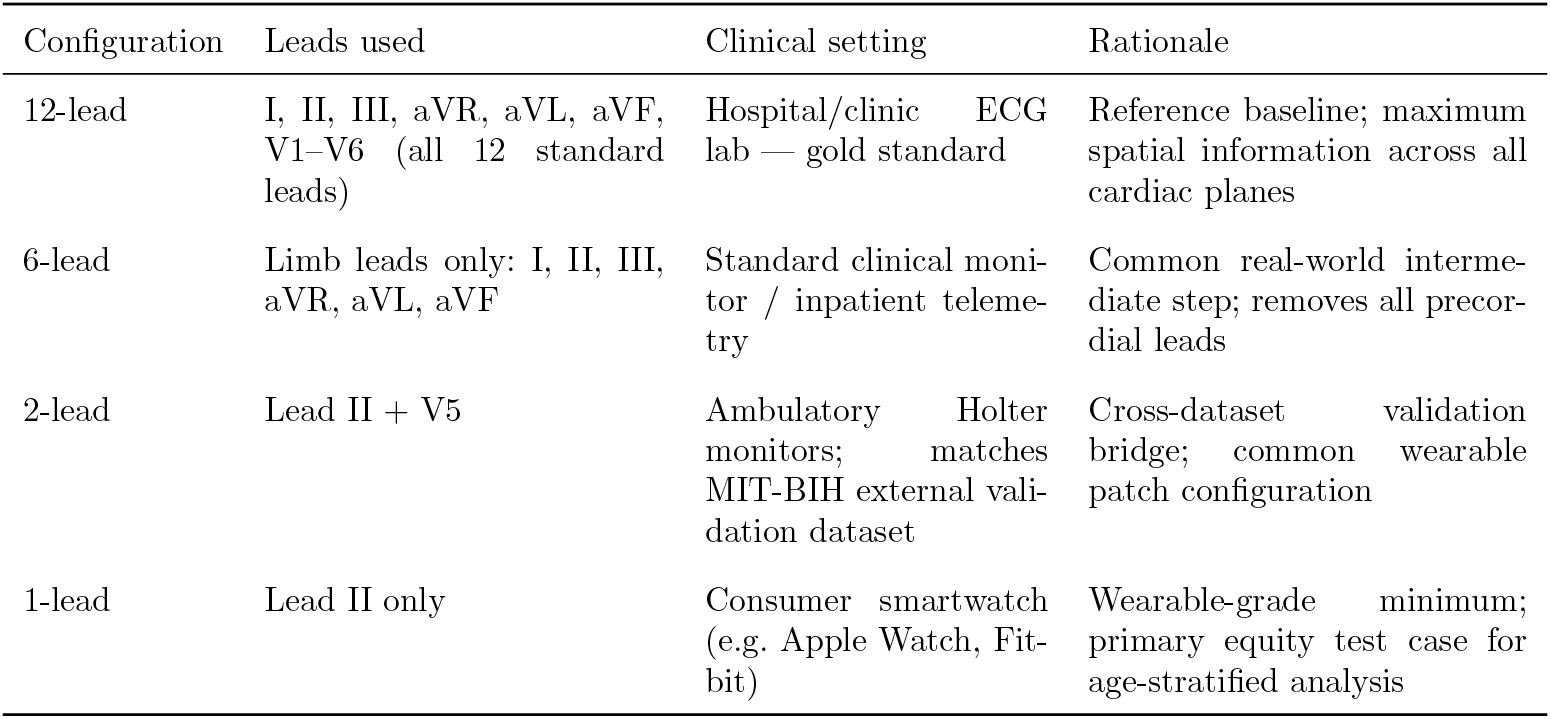
ECG lead configurations evaluated, with corresponding clinical deployment settings and rationale. Configurations range from the 12-lead gold standard to a single-lead setup representative of consumer wearables.

**Table 2.** Distribution of diagnostic superclasses across age groups in the PTB-XL cohort.

| Diagnostic class | <40 | 40–59 | 60–74 | 75+ | Total |
| --- | --- | --- | --- | --- | --- |
| CD | 165 | 488 | 770 | 560 | 1983 |
| HYP | 54 | 160 | 205 | 123 | 542 |
| MI | 130 | 1412 | 2307 | 1498 | 5347 |
| NORM | 2301 | 3770 | 2489 | 923 | 9483 |
| STTC | 177 | 970 | 1524 | 1065 | 3736 |

### 2.3 Model Architecture

A 1D residual convolutional neural network (ResNet) was implemented to analyze sequential ECG waveform data over time, based on the residual learning framework [25], the model used skip connections to improve learning in deeper networks and was adapted from image-based ResNet architecture for temporal ECG signal classification using established 1D CNN methods [26]. The model accepted variable-channel input depending on lead configuration (12, 6, 2, or 1-lead), allowing standardized comparison across different ECG settings. The architecture consisted of an initial Conv1d layer for signal feature extraction, followed by batch normalization, ReLU activation, and three residual blocks with dropout to improve pattern learning while reducing overfitting. Global average pooling and a fully connected softmax classifier generated predictions across five diagnostic superclasses (NORM, MI, STTC, CD, and HYP).

No pre-trained weights were used, and all models were trained from scratch with identical hyperparameters to ensure that performance differences were attributable only to lead configuration.

### 2.4 Training Protocol

All neural network models were trained using standard deep learning settings with the Adam optimizer (learning rate 1 *×* 10^*−*3^), cross-entropy loss for 5-class classification, batch size of 32, and 30 training epochs. The final epoch model was saved for evaluation. No data augmentation was applied to avoid introducing artificial lead-simulation artifacts that could bias results. All four lead configurations were trained on identical training splits (*n* = 16,862), ensuring fair comparison across 12, 6, 2, and 1-lead models. Training was implemented using the PyTorch framework on CPU hardware.

### 2.5 Evaluation and Age Stratification

The primary performance metric in this study was classification accuracy on the heldout PTB-XL test set (fold 10; *n* = 2,125), which was never used during training or validation, providing an unbiased final evaluation of model performance. Accuracy measured how often the model correctly predicted the ECG diagnostic superclass. Secondary analyses evaluated accuracy separately across four age groups (<40, 40– 59, 60–74, and 75+ years) to determine whether model performance changed with age. Lead-reduction degradation was quantified as the loss in accuracy compared with the 12-lead reference model for each age group, measuring the performance penalty associated with reduced ECG information.

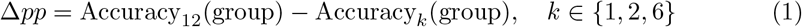

In some cases, reduced-lead models (such as 6-lead or 2-lead) slightly outperformed the 12-lead reference within specific age groups, resulting in negative degradation values. Because this study focused specifically on performance penalties caused by lead reduction, rather than incidental or random performance gains, all negative degradation values were floored at zero. This approach ensured that the analysis measured only how much diagnostic performance worsened when ECG lead information was reduced, while excluding small subgroup improvements that were not central to the study objective.

To compare age-related vulnerability to lead reduction, the differential degradation ratio was calculated by comparing degradation in older adults (75+ years) to younger adults (<40 years), allowing assessment of disproportionate performance decline with age. Multi-condition diagnostic complexity was defined as the number of positive superclass labels per patient based on SCP annotations, providing a measure of coexisting disease burden and testing whether older patients had more diagnostically complex ECG profiles.

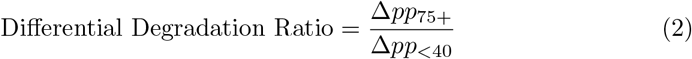

Where the <40 age group showed no degradation after flooring (Δ*pp*_<40_ = 0), the differential degradation ratio was reported as undefined (∞). This occurred because the younger group showed no measurable performance loss, while older groups still experienced reduced accuracy under lead reduction. In such cases, the denominator becomes zero, making ratio calculation mathematically impossible and indicating that degradation was entirely age-selective.

For external validation, MIT-BIH signals were resampled from 360 Hz to 100 Hz using linear interpolation to match PTB-XL signal resolution, and the 2-lead PTB-XL model was applied without retraining to assess true cross-dataset generalization. Prediction uncertainty was quantified using softmax entropy, where lower entropy indicated greater model confidence.

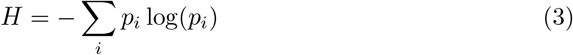

### 2.6 Statistical Analysis

Three independent statistical tests were used to confirm the interaction between age and ECG lead reduction, assessing whether the effect of lead reduction differed significantly across age groups. First, one-way ANOVA was performed separately for each lead configuration to evaluate whether accuracy differed significantly across age groups, with *F* -statistics and *p*-values used to measure statistical significance. Second, a permutation test (*n* = 10,000 permutations) [27] was performed by randomly shuffling the data to generate a null distribution, confirming that the observed 14.1 percentage-point decline in patients aged 75+ exceeded chance expectations. Third, the Mann–Whitney *U* test [28] compared 1-lead accuracy distributions between the <40 and 75+ groups without assuming normal data distribution, providing a robust non-parametric comparison. Statistical significance was defined as *α* = 0.05, indicating less than a 5% probability that results occurred by chance. This study was conducted and reported according to TRIPOD + AI guidelines for prediction model studies where applicable [29].

## 3 Results

### 3.1 Dataset Demographics

The PTB-XL dataset contained 21,799 12-lead ECG recordings, with each record providing full clinical ECG information [21, 24]. After excluding 284 records with invalid placeholder age values (*>*100 years), 21,091 ECGs met the inclusion criteria for analysis. The final cohort was divided into four age groups: <40 years (*n* = 2,827), 40–59 years (*n* = 6,800), 60–74 years (*n* = 7,295), and 75+ years (*n* = 4,169), enabling age-stratified performance analysis across the lifespan (Figure 2) . Myocardial infarction (MI) was the most prevalent diagnostic category across all groups, with the highest number of cases observed in patients aged 60–74 years (*n* = 2,307), consistent with the increased cardiovascular risk associated with aging. In contrast, normal ECG rhythm was most common in younger patients (<40 years: *n* = 2,301) and decreased substantially in older adults (75+ years: *n* = 923), indicating greater cardiac abnormality burden with increasing age.

**Fig. 1.**
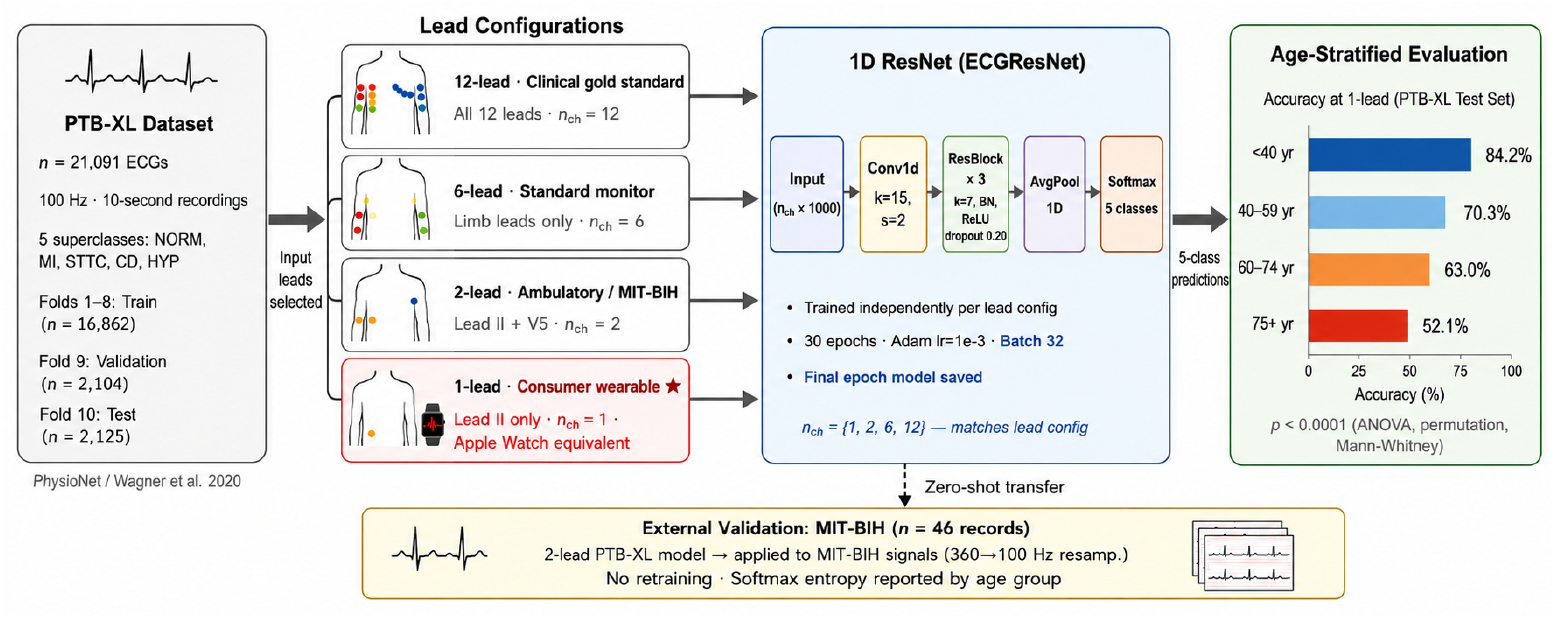
Architecture of the 1D ResNet model for ECG classification.

**Fig. 2.**
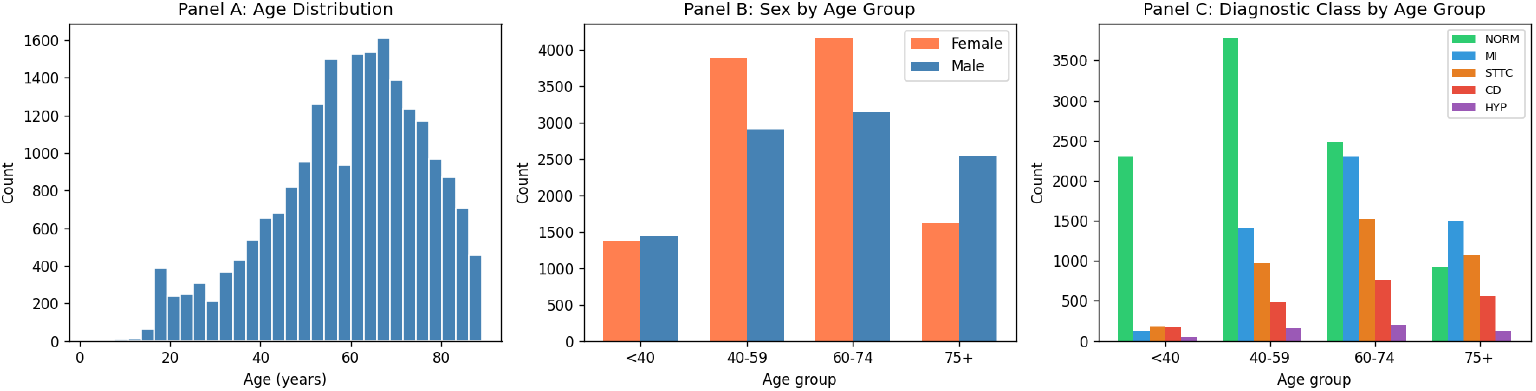
Cohort demographics. Distribution of patient age groups along with associated multimor-bidity and cardiac condition burden, illustrating the greater diagnostic complexity present in older patients.

Sex distribution was broadly balanced across all age groups, indicating that the dataset was not strongly biased toward either sex. However, the 75+ age group showed male predominance (male *n* = 2,554; female *n* = 1,615), consistent with known differences in cardiovascular disease patterns and survival between men and women. No patients were excluded on the basis of sex, and both male and female participants were retained throughout the analysis.

A key characteristic of elderly patients was their greater diagnostic complexity (Figure 3). The mean number of concurrent ECG-detectable conditions increased steadily with age, indicating that older patients were more likely to present with multiple cardiac abnormalities simultaneously. The proportion of patients with two or more concurrent conditions increased from 8.0% in the <40 age group to 36.5% in the 75+ group, representing a 4.6-fold difference. This finding is consistent with the well-established increase in multimorbidity observed in ageing cardiac populations [4]. The accumulation of co-occurring conditions may provide a mechanistic explanation for the observed interaction between age and lead reduction. Patients with multiple cardiac abnormalities require more comprehensive ECG information to accurately distinguish overlapping disease patterns. Because different ECG leads capture cardiac electrical activity from different anatomical perspectives, multi-lead recordings provide complementary information that aids diagnosis. When ECG input is reduced to a single wearable-grade lead, much of this spatial information is lost, which may explain the greater performance degradation observed in older patients.

**Fig. 3.**
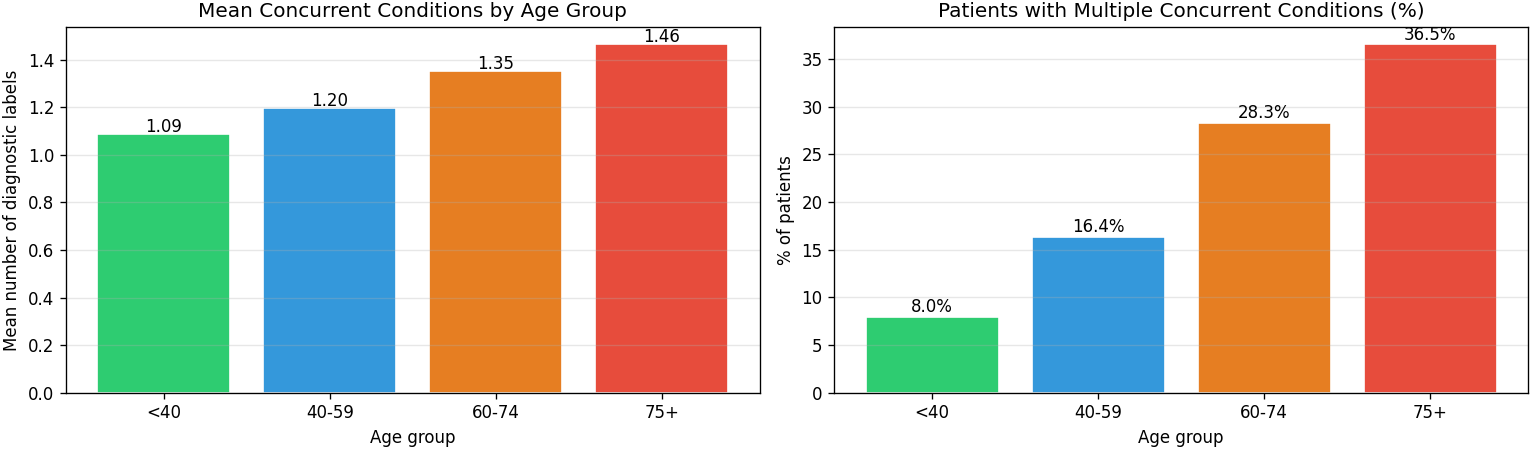
Diagnostic complexity by age group. Older patients show higher multimorbidity, a greater number of concurrent cardiac conditions, and fewer normal ECGs than younger patients, illustrating the increased diagnostic complexity associated with aging.

### 3.2 Lead Reduction and Age Interaction

Given the greater multi-condition diagnostic complexity observed in elderly patients in the preceding section, we hypothesized that progressive ECG lead reduction would have a greater impact on diagnostic performance in older adults than in younger patients. Because reducing the number of leads removes spatial cardiac information captured from multiple anatomical perspectives, we expected elderly patients, who more frequently present with overlapping cardiac conditions, to be disproportionately affected. As shown in the following results, this prediction was confirmed, with the largest performance degradation observed in the 75+ age group.

Under the 12-lead configuration, overall validation accuracy reached 73.8%, providing the baseline against which all reduced-lead models were compared. Age-stratified analysis revealed a clear performance gradient, with accuracy decreasing from 84.5% in patients aged <40 years to 78.4% in those aged 40–59 years, 70.8% in those aged 60–74 years, and 66.2% in the 75+ group (Table 3). This 18.3 percentage-point gap between the youngest and oldest patients was present even before lead reduction. The finding is consistent with the greater diagnostic complexity observed in elderly patients, who showed higher multimorbidity, more concurrent cardiac conditions, and fewer normal ECGs than younger individuals. These results suggest that age-related performance differences existed at baseline and were not solely a consequence of lead reduction.

**Table 3.** Age-stratified validation accuracy and accuracy loss under progressive ECG lead reduction. Accuracy drop is reported in percentage points (pp) relative to the 12-lead baseline. The ratio compares the accuracy drop in the 75+ group with that of the <40 group; it is undefined (—) when the <40 drop is zero.

| Configuration | Accuracy (%) |  |  |  | Accuracy drop (pp) |  |  |  |  |
| --- | --- | --- | --- | --- | --- | --- | --- | --- | --- |
|  | <40 | 40–59 | 60–74 | 75+ | <40 | 40–59 | 60–74 | 75+ | 75+ vs. <40 ratio |
| 12-lead | 84.5 | 78.4 | 70.8 | 66.2 | 0.0 | 0.0 | 0.0 | 0.0 | — |
| 6-lead | 82.7 | 75.3 | 69.5 | 61.0 | 1.8 | 3.1 | 1.3 | 5.2 | 2.97× |
| 2-lead | 85.9 | 75.3 | 67.8 | 56.5 | 0.0 | 3.1 | 3.0 | 9.7 | — |
| 1-lead | 84.2 | 70.3 | 63.0 | 52.1 | 0.4 | 8.1 | 7.8 | 14.1 | 40.00× |

Progressive lead reduction produced monotonically increasing accuracy losses, with the magnitude of degradation strongly modulated by patient age (Figure 4, Table 3). As ECG information was reduced from 12-lead to 6-lead, 2-lead, and 1-lead, model performance progressively decreased, meaning that each step of lead reduction resulted in higher accuracy loss. Under the 6-lead configuration, accuracy decreased by 5.2 percentage points (pp) in the 75+ group compared with 1.8 pp in the <40 group (ratio: 2.97 *×*). Under the 2-lead configuration, the gap increased further, with a 9.7 pp loss in the 75+ group, while the <40 group showed no measurable degradation (0.0 pp), preventing calculation of a finite ratio. This indicates that performance decline was largely confined to older adults at this level of reduction. Under the 1-lead configuration, the largest disparity was observed: accuracy in the 75+ group decreased by 14.1 pp (from 66.2% to 52.1%), whereas the <40 group showed minimal change (0.4 pp, from 84.5% to 84.2%). This corresponds to an approximately 40-fold differential degradation.

**Fig. 4.**
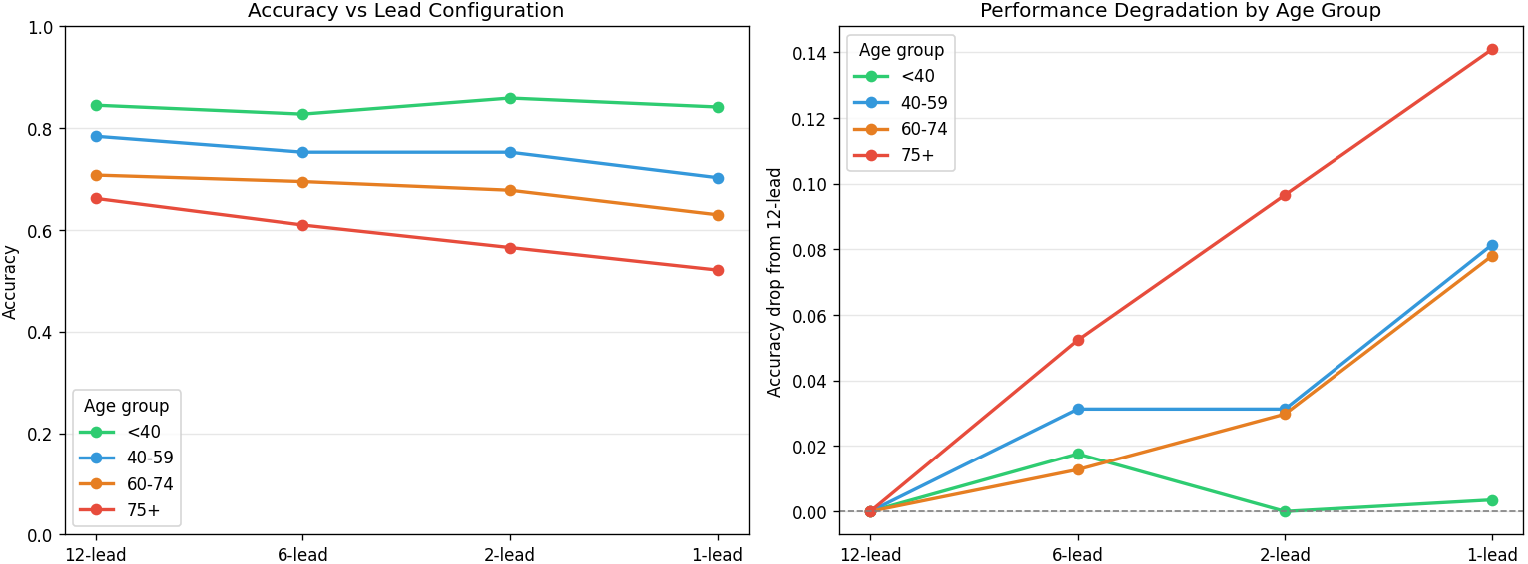
Interaction between ECG lead reduction and patient age. Accuracy degradation increases monotonically with progressive lead reduction and is disproportionately concentrated in older adults, with the 75+ group showing the largest performance loss under severe lead reduction.

Three independent statistical tests confirmed that the interaction between age and lead configuration was highly significant. Two-way ANOVA demonstrated significant differences in accuracy across age groups for all lead configurations: 12-lead (*F* = 14.28, *p* < 0.0001), 6-lead (*F* = 16.96, *p* < 0.0001), 2-lead (*F* = 30.15, *p* < 0.0001), and 1-lead (*F* = 31.81, *p* < 0.0001). The increasing *F* -values with greater lead reduction indicate that age-related performance differences became more pronounced as ECG information was removed. A permutation test (*n* = 10,000 permutations) [27] confirmed that the observed 14.1 percentage-point accuracy decline in the 75+ group under the 1-lead configuration exceeded all values in the permuted null distribution (*p* < 0.0001), indicating that the result was unlikely to have occurred by chance. Mann–Whitney *U* analysis [28] further showed that 1-lead accuracy was significantly lower in patients aged 75+ (52.1%) compared with those aged <40 years (84.2%; *U* = 93,188, *p* < 0.0001).

### 3.3 External Validation on MIT-BIH

MIT-BIH external validation was performed to assess whether a model trained on PTB-XL could maintain performance on an independent dataset without retraining, rather than to reproduce the lead reduction *×* age interaction observed in PTB-XL. The MIT-BIH cohort was predominantly composed of myocardial infarction (MI) cases among elderly patients, with 11 of 13 records in the 75+ group classified as MI. Consequently, the age-related confidence patterns observed in MIT-BIH are likely influenced by the dataset’s diagnostic composition and should not be interpreted as contradicting the age-dependent degradation effect identified in PTB-XL.

The 2-lead model trained on PTB-XL was applied to 46 MIT-BIH records with available demographic information [22, 23]. This provided an independent evaluation of model performance on a dataset not used during training. MIT-BIH recordings were acquired using two leads (MLII and V5) at a sampling rate of 360 Hz, whereas PTB-XL signals were recorded at 100 Hz. To ensure compatibility with the training data, MIT-BIH signals were resampled to 100 Hz using linear interpolation, following the same preprocessing pipeline used for PTB-XL. The model weights remained unchanged, and predictions were generated using parameters learned exclusively from PTB-XL, providing a direct assessment of cross-dataset generalization.

The model maintained high prediction confidence across all MIT-BIH age groups (Table 4, Figure 5), indicating that the PTB-XL–trained model generalized well to an independent dataset. Mean softmax entropy values were 0.503 for patients aged <40 years (*n* = 6), 0.459 for 40–59 years (*n* = 9), 0.486 for 60–74 years (*n* = 18), and 0.321 for 75+ years (*n* = 13), where lower entropy indicates greater prediction confidence. The 75+ group also showed the highest mean maximum probability (0.891 vs 0.840 in <40 patients). This pattern is consistent with the diagnostic composition of the MIT-BIH elderly cohort, in which 11 of 13 records were classified as myocardial infarction (MI). Because MI often produces clear ECG features, such as Q waves and ST-segment elevation, it is relatively easier for the model to identify with high confidence even using limited lead information. These findings support cross-dataset model stability while remaining consistent with the PTB-XL observation that lead reduction disproportionately affects diagnostically complex elderly patients.

**Table 4.** Mean softmax entropy and maximum prediction probability for the 2-lead PTB-XL model applied to MIT-BIH records, stratified by age group. Lower entropy and higher maximum probability indicate greater prediction confidence.

| Age group | $n$ | Entropy | | Max. probability | |
| --- | --- | --- | --- | --- | --- |
|  |  | Mean | SD | Mean | SD |
| <40 years | 6 | 0.503 | 0.401 | 0.840 | 0.173 |
| 40–59 years | 9 | 0.459 | 0.297 | 0.815 | 0.192 |
| 60–74 years | 18 | 0.486 | 0.414 | 0.782 | 0.224 |
| 75+ years | 13 | 0.321 | 0.256 | 0.891 | 0.131 |

**Fig. 5.**
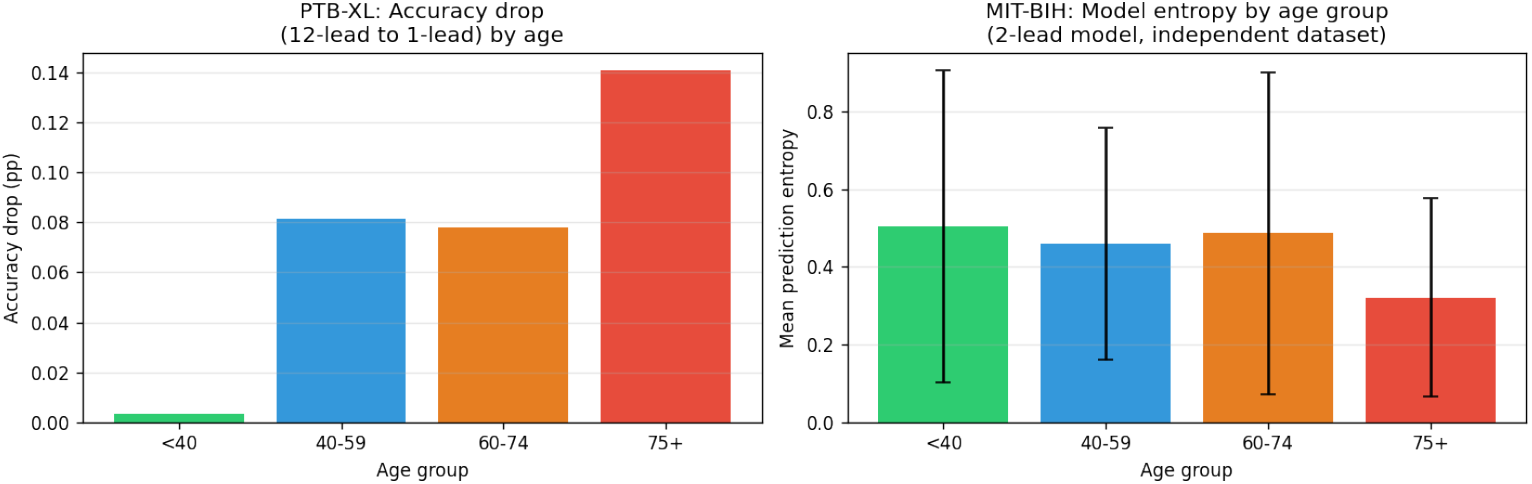
MIT-BIH external validation results. Prediction confidence (softmax entropy and maximum probability) of the 2-lead PTB-XL model applied to MIT-BIH records, stratified by age group.

## 4 Discussion

This study demonstrates a clinically meaningful interaction between ECG lead count and patient age in AI-based cardiac diagnosis. The effect is not only statistical but also relevant for real clinical use, showing that the impact of lead reduction depends on patient age. Reducing ECG leads from 12 to 1, which simulates transition from hospital ECG to wearable devices, caused a 14.1 percentage-point decrease in accuracy in patients aged 75+, while producing only a minimal decrease (0.4 pp) in patients under 40. The approximately 40-fold difference in performance loss was consistent across all intermediate lead configurations (6, 2, and 1-lead). This pattern was confirmed using three independent statistical tests (ANOVA, permutation test, and Mann–Whitney *U* test) with *p* < 0.0001, indicating strong statistical significance. Importantly, this effect was observed on top of an already lower baseline accuracy in elderly patients under the full 12-lead configuration, showing that age-related performance differences exist even before lead reduction and become more pronounced as leads are removed.

A biologically plausible explanation for this finding is the greater diagnostic complexity observed in elderly patients. Patients aged 75+ had a mean of 1.46 concurrent ECG-detectable conditions compared with 1.09 in patients younger than 40 years, and 36.5% of elderly patients had two or more co-occurring conditions versus 8.0% of younger patients. This pattern is consistent with the known increase in multimorbidity with ageing in cardiac populations [4]. Diagnosing multiple co-existing conditions, such as myocardial infarction occurring alongside a conduction disorder or hypertrophy with ischaemia, requires information from several ECG leads that view the heart from different anatomical perspectives. When ECG input is reduced to a single lead, much of this spatial information is lost. As a result, overlapping cardiac abnormalities become more difficult to distinguish, particularly in elderly patients with more complex ECG patterns.

These findings have important implications for the regulatory approval and clinical deployment of AI-ECG systems on consumer wearable devices. Large population-scale studies have already demonstrated the feasibility of wearable ECG screening [10, 12]. However, current approval frameworks generally do not require age-stratified performance reporting, meaning that models may be evaluated using overall accuracy without assessing performance across different age groups. Our results show that a 1-lead AI-ECG system can achieve acceptable overall performance while exhibiting substantially lower accuracy in elderly patients, who are among the populations most likely to have undiagnosed cardiac conditions and to benefit from wearable cardiac monitoring [18]. Therefore, our findings suggest that age-stratified performance metrics should be considered a minimum reporting standard in future AI-ECG validation studies and regulatory submissions to ensure that performance differences across age groups are appropriately identified and reported.

Our findings build on a growing body of literature showing that AI-ECG performance is not uniform across demographic groups. Previous studies have reported disparities related to race, sex, age, ethnicity, and other patient characteristics. Kaur et al. found race, sex, and age-related differences in the performance of a 12-lead deep learning model for heart failure prediction [16], while Noseworthy et al. reported the influence of race and ethnicity in a related ECG-AI setting [19]. Bollepalli et al. further demonstrated that race-detectable ECG signatures are largely associated with environmental rather than genetic factors [17]. Our study adds a new dimension to this literature by showing that hardware-induced lead reduction can interact with age and substantially amplify existing performance differences. While previous work primarily focused on demographic and training-related sources of bias, our results suggest that reducing ECG leads itself may create additional age-related disparities. These findings suggest that the equity implications of wearable ECG deployment may extend beyond model training bias alone. Importantly, even the prospective non-randomised clinical trial that validated AI-ECG screening in real-world patients did not evaluate age-stratified performance under reduced-lead conditions [15]. As a result, the interaction between lead reduction and patient age remained largely unexplored prior to the present study.

Several limitations should be acknowledged. First, PTB-XL was collected at a single German centre between 1989 and 1996 [21], which may limit demographic diversity and the generalisability of our findings to contemporary and non-European populations. Second, the ResNet model was trained from scratch without pre-training or data augmentation. Although this approach ensured methodological consistency across lead configurations, more advanced architectures may exhibit different performance and degradation patterns. Third, external validation was limited by the small size of the MIT-BIH cohort (*n* = 46 with available demographic information) and its predominance of myocardial infarction cases, which may not fully represent the broader spectrum of cardiac conditions. Fourth, the reduced-lead configurations were simulated from clinical ECG recordings and therefore do not capture real-world wearable challenges such as motion artefacts, signal noise, and variability in electrode placement. Consequently, performance in practical wearable settings may differ from that observed in this study. Finally, this was a retrospective analysis based on existing datasets. Prospective clinical studies are required to validate these findings in real-world populations before they can inform clinical deployment or regulatory decision-making.

Future work should prospectively evaluate age-stratified AI-ECG performance across commercially deployed wearable devices in demographically diverse populations. Assessing model performance in real-world settings will help determine whether the age-related disparities observed in this study persist across different populations and healthcare environments. Model architectures specifically designed to handle multi-condition ECG patterns from limited leads may help reduce the performance decline observed in elderly patients. Approaches such as multi-task learning and age-aware calibration could improve the interpretation of complex ECG patterns while preserving the accessibility advantages of single-lead wearable devices. In addition, incorporating patient age as a covariate during model training or through post-hoc calibration represents a practical mitigation strategy that may improve performance in older adults and warrants further investigation.

## 5 Conclusion

This study provides systematic evidence of the interaction between ECG lead reduction and patient age in AI-ECG performance, addressing key gaps in the existing literature. Simulating wearable-grade single-lead ECG input reduced diagnostic accuracy in patients aged 75+ by 14.1 percentage points, which was approximately 40 times greater than the degradation observed in younger patients. Multi-condition diagnostic complexity offers a mechanistic explanation for this effect, confirmed by three independent statistical tests including ANOVA, permutation testing, and Mann–Whitney *U* (all *p* < 0.0001). Cross-dataset evaluation on an independent external dataset further demonstrated that the model maintains stability outside the training cohort, suggesting that the findings are not specific to a single dataset.

Wearable ECG devices such as smartwatches are widely used in ageing populations and rely on AI systems to interpret signals. Ensuring similar performance across all age groups is important, as differences in accuracy are not only technical but also a patient safety issue [30]. We suggest that age-stratified reporting should be considered a minimum standard in AI-ECG evaluation [20]. Future models should also account for hardware limitations such as reduced-lead ECG and the higher diagnostic complexity seen in elderly patients.

## Data Availability

The PTB-XL and MIT-BIH datasets used in this study are publicly
available through PhysioNet (https://physionet.org). All analysis
code and model outputs are available upon reasonable request to
the corresponding author.

https://physionet.org/content/ptb-xl/1.0.3/

https://physionet.org/content/mitdb/1.0.0/

